# Ethnoracial Disparities in Breast Cancer Treatment Time and Survival: A Systematic Review With a DAG-based Causal Model

**DOI:** 10.1101/2024.06.02.24308338

**Authors:** Parisa M. Hesari, Drexler James, Daniel J. Lizotte, Greta R. Bauer

**Author notes:** Correspondent authors’ Email, address, and, Address: Schulich School of Medicine & Dentistry, Western University, Epidemiology & Biostatistics, 3rd Floor, 1465 Richmond St, London, ON, Canada N6G 2M1 t., www.westernu.ca.

## Abstract

For interventions aimed at redressing health disparities in breast cancer to be effective, a clear understanding of the nature and causes of these disparities is required. Our question is: what is the current evidence for ethnoracial disparities in time-to-treatment initiation and survival in breast cancer, and how are the causal mechanisms of these disparities conceptualized in the literature? A comprehensive systematic search of studies on cohorts of female breast cancer patients diagnosed with stage I-III was performed. Directed acyclic graphs were used to describe implicit causal relationships between ethnoracial group membership and time-to-treatment initiation and survival outcomes. This review revealed strong evidence for ethnoracial disparities in both time to treatment and survival among breast cancer patients. Unmeasured factors identified by the authors highlighted gaps in data sources and opportunities for causal reasoning. While the existing literature describes ethnoracial disparities, there is very limited discussion of causal mechanisms, and no discussion of system-level rather than individual-level effects. In response, a biosocioecological model of breast cancer disparity was developed to integrate system-level considerations into future research. Addressing established ethnoracial disparities in breast cancer requires new research that explicitly considers the causal mechanisms of potential interventions, incorporating unmeasured factors contributing to these disparities.

## Background

Inequalities in access to care across ethnoracial groups are a known driver of differential health outcomes [1]. An increasing body of evidence demonstrates that communities of color have been disproportionately affected by systemic racism that leads to disparities in chronic disease outcomes, including for breast cancer [2–4]. Though breast cancer survival has increased globally due to advances in survivorship care, treatment modalities, and prevention methods [4,5], studies have shown less improved survival in marginalized groups, even after controlling for biologic and other characteristics [6,7].

There is increasing evidence that factors associated with delays in treatment initiation vary across ethnoracial groups of breast cancer patients, and that this drives disparities in outcomes [8–10]. Studies examining the impact of therapy duration on survival across these groups have identified significant differences in time to treatment initiation and survival [10,11]. Many factors affecting disparities in breast cancer have been identified, including social and health determinants along with individualized and tumor-related factors [12–16]. However, many studies have prioritized establishing association rather than examining the causal mechanisms underlying these associations [17,18], and many studies rely on cancer registry data, which often do not contain the information needed to assess such mechanisms.

Causal reasoning tools, such as directed acyclic graphs (DAGs), can enable researchers and policymakers to clarify and assess potential causal mechanisms. Understanding mechanisms that influence time to treatment initiation has the potential to inform targeted individual and system-level interventions to mitigate ethnoracial disparities and improve health outcomes [19].

Despite this potential value, there has been to date no overarching synthesis of causal reasoning on ethnoracial disparities in treatment initiation in breast cancer. This study represents a pioneering effort to systematically review the causal logic of the existing published evidence that assesses disparities in Time to Treatment Initiation (TTI) and survival across ethnoracial groups. To achieve this, we construct and combine study-specific DAGs, producing a summary DAG showing the structure of causal logic used within the field to describe relationships among ethnoraciality and other factors and their relationship to TTI and breast cancer survival. The resulting knowledge allows us to both 1) describe key causal logic structures considered in existing literature, and 2) identify individual and structural factors that are known to impact access to care or ethnoracial disparities, but have not been addressed in the context of breast cancer.

## Methods

This systematic review is registered in the PROSPERO International Prospective Register of Systematic Reviews (ID: CRD42023391901). Our methodology included two primary activities. First, we systematically reviewed literature on ethnoracial disparities and time to treatment and/or survival, adhering to the Preferred Reporting Items for Systematic Review and Meta-Analysis Protocols (PRISMA-P) [20] guidelines using the Covidence platform [21]. Second, we constructed DAGs representing the presumed causal relationships among factors in each included study, and consolidated them into a single DAG, following a modification of the evidence synthesis for constructing directed acyclic graphs (ESC-DAGs) guideline [22].

### i. Systematic literature review on ethnoracial disparities

#### Eligibility criteria

English language peer-reviewed cohort studies published in academic journals were eligible for inclusion if they focused on patients aged 18 years and above diagnosed with stage I to III breast cancer between 1995 and 2019. Study participants must have undergone at least one form of systemic or local therapy.

Studies that failed to consider race/ethnicity were excluded. Studies that did not explicitly exclude stage IV breast cancer patients were excluded, as treatment primarily involves palliative care. Studies during the COVID-19 pandemic were excluded due to pandemic-related changes in the treatment modalities and management of breast cancer patients, influenced by both individual and healthcare system factors.

#### Outcomes: Time to treatment and survival measure

We considered two outcome measures: time to treatment initiation (TTI) and survival. TTI was defined as the time from diagnosis to initiation of local therapies (e.g., surgery) and/or systemic therapies (e.g., chemotherapy.) Survival time was measured as the interval between treatment initiation and events like breast cancer-related deaths or follow-up time for survivors.

#### Search strategy

Eligible studies were identified in the PubMed, Ovid, Web of Science, and Cochrane Library databases. A title-filtered search was also conducted through Google Scholar using the keywords ’breast cancer’ and ’time to treatment’, as was a manual search using reference lists of the included studies. Primary keywords ’breast cancer’ and ’time to treatment’ were searched (eTable1 in Supplement1). We did not require that ’race’ appear in the title or abstract, as this would exclude many relevant studies. Full details are provided in the protocol document [23].

#### Study records

Duplicate records were removed, the remaining studies were screened based on title and abstract, and then full texts were assessed for inclusion. Data extraction was performed by two independent reviewers and reviewers convened to discuss any uncertainties, including any errors in data collection process or uncertainty regarding inclusion of a study. Studies with correction or retraction notices were removed.

#### Data extraction and Risk of bias assessment

We extracted study characteristics including main objective, study design, source of data, sample size, data collection period, ethnoracial group categories, types of treatments assessed, and lists of author-identified unmeasured factors, using Covidence. Two reviewers assessed risk of bias using a tool developed by the CLARITY group at McMaster University [24] to categorize each study as having low, moderate, or high risk of bias. In the event of discrepancies, a third reviewer was consulted. We updated the search by March 2024.

### ii. Construction of DAGs

To synthesize causal relationships among identified factors within the included studies, we constructed: (1) A *study-specific DAG* for each study individually (eFigure 1 to eFigure 36 in Supplement 1), and (2) a single *comprehensive DAG* that combined factors across all studies.

**Figure 1.**
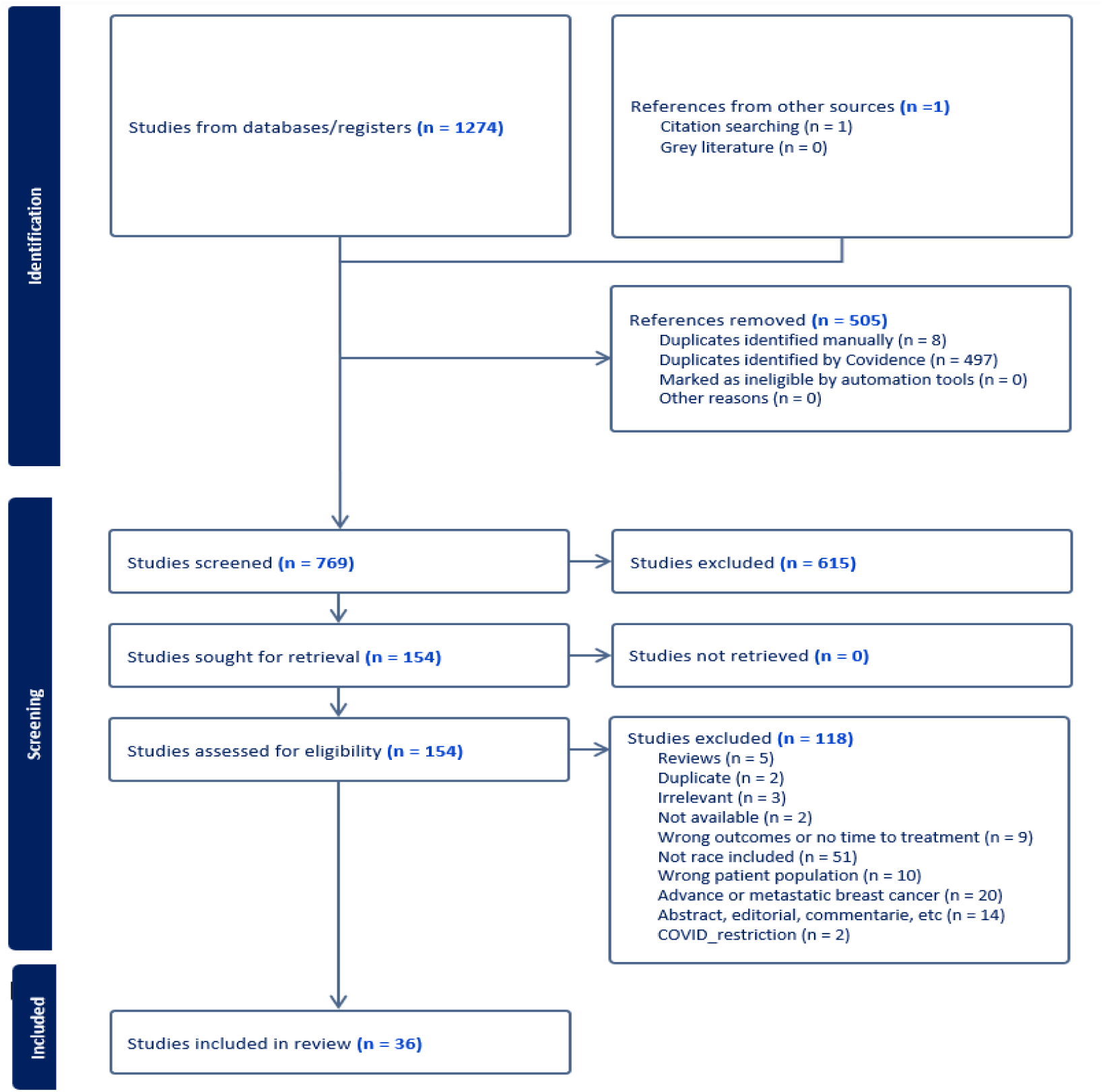
Process of identification and data extraction of studies on racial disparity and time to treatment on survival in breast cancer using PRISMA.

#### Study-specific DAGs

To construct *study-specific DAGs*, we systematically extracted factors investigated by study authors, recognizing that most of these factors were not explicitly evaluated for causal relationships. We aimed for *study-specific DAGs* to describe the literature (as most of that is not explicitly causal,) identify implicit causal models, and integrate causal reasoning.

#### Producing the Comprehensive DAG

The comprehensive DAG was constructed based on the observed relationships between ethnoracial group membership (EGM), other variables, and the outcomes of interest: TTI and survival. To manage the large number of other variables assessed in the literature, we grouped variables into a single node if 1) there was face validity in combining them into a broader construct, and 2) if they had the same parent nodes and same child nodes, and 3) if they had similar impact and hypothesized direction along relevant causal pathways. We considered effects along two major causal pathways: 1) the direct causal pathway(s) from EGM to TTI and/or survival, and 2) in models that consider a mediator, the indirect causal pathway(s) related to the mediator. For instance, we would only combine income, education, and occupation into a single socioeconomic status (SES) node if their effects on relevant causal pathways align in the same direction (positive or negative) and if they had the same parents and children.

## Results

We initially identified 1,346 records (Figure 1) which, after removing duplicates and screening, yielded 151 studies for full-text review. Of these, 36 were included in the final analysis. All factors assessed in the literature are summarized in Table 1, grouped according to the principles described above.

**Table 1.**
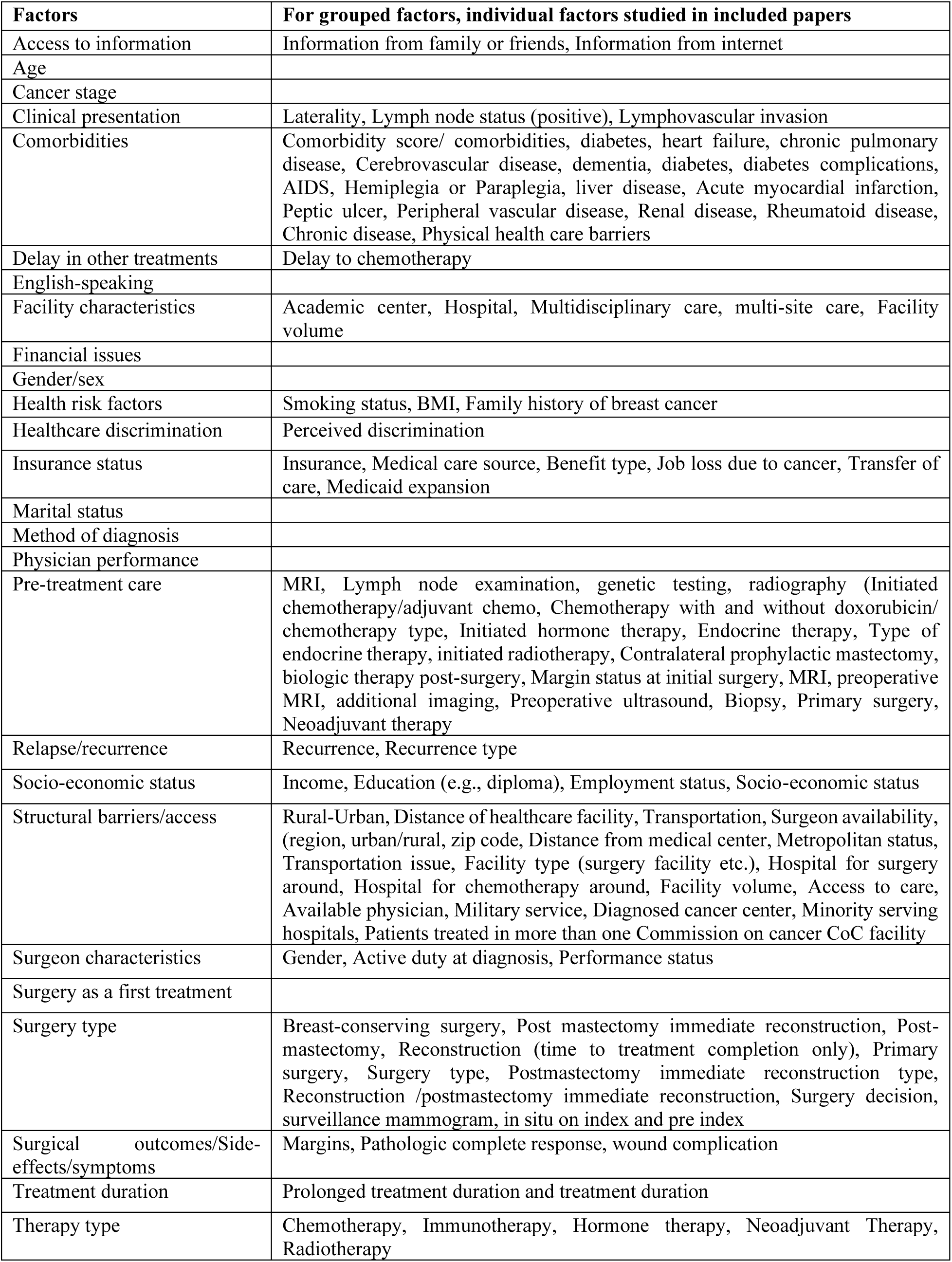
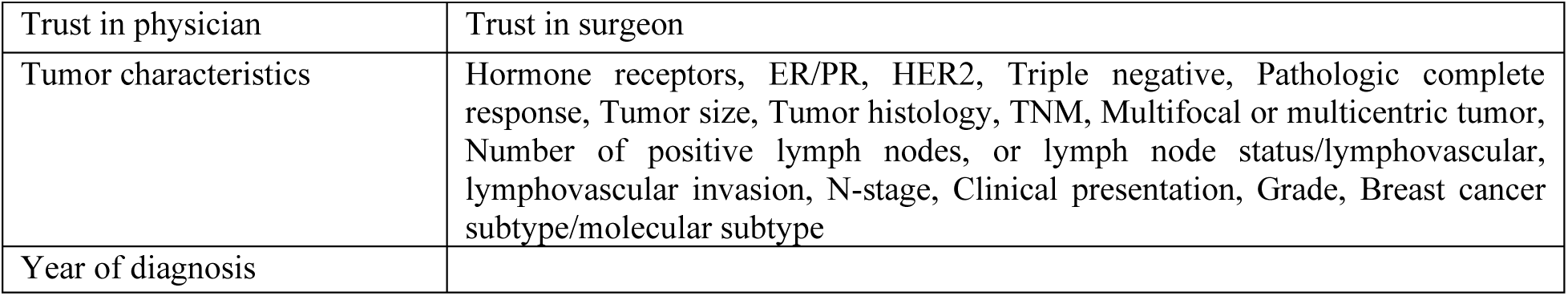
List of factors from included studies and detailed factors within each group.

| **Factors** | **For grouped factors, individual factors studied in included papers** |
| --- | --- |
| Access to information | Information from family or friends, Information from internet |
| Age |  |
| Cancer stage |  |
| Clinical presentation | Laterality, Lymph node status (positive), Lymphovascular invasion |
| Comorbidities | Comorbidity score/ comorbidities, diabetes, heart failure, chronic pulmonary disease, Cerebrovascular disease, dementia, diabetes, diabetes complications, AIDS, Hemiplegia or Paraplegia, liver disease, Acute myocardial infarction, Peptic ulcer, Peripheral vascular disease, Renal disease, Rheumatoid disease, Chronic disease, Physical health care barriers |
| Delay in other treatments | Delay to chemotherapy |
| English-speaking |  |
| Facility characteristics | Academic center, Hospital, Multidisciplinary care, multi-site care, Facility volume |
| Financial issues |  |
| Gender/sex |  |
| Health risk factors | Smoking status, BMI, Family history of breast cancer |
| Healthcare discrimination | Perceived discrimination |
| Insurance status | Insurance, Medical care source, Benefit type, Job loss due to cancer, Transfer of care, Medicaid expansion |
| Marital status |  |
| Method of diagnosis |  |
| Physician performance |  |
| Pre-treatment care | MRI, Lymph node examination, genetic testing, radiography (Initiated chemotherapy/adjuvant chemo, Chemotherapy with and without doxorubicin/ chemotherapy type, Initiated hormone therapy, Endocrine therapy, Type of endocrine therapy, initiated radiotherapy, Contralateral prophylactic mastectomy, biologic therapy post-surgery, Margin status at initial surgery, MRI, preoperative MRI, additional imaging, Preoperative ultrasound, Biopsy, Primary surgery, Neoadjuvant therapy |
| Relapse/recurrence | Recurrence, Recurrence type |
| Socio-economic status | Income, Education (e.g., diploma), Employment status, Socio-economic status |
| Structural barriers/access | Rural-Urban, Distance of healthcare facility, Transportation, Surgeon availability, (region, urban/rural, zip code, Distance from medical center, Metropolitan status, Transportation issue, Facility type (surgery facility etc.), Hospital for surgery around, Hospital for chemotherapy around, Facility volume, Access to care, Available physician, Military service, Diagnosed cancer center, Minority serving hospitals, Patients treated in more than one Commission on cancer CoC facility |
| Surgeon characteristics | Gender, Active duty at diagnosis, Performance status |
| Surgery as a first treatment |  |
| Surgery type | Breast-conserving surgery, Post mastectomy immediate reconstruction, Post-mastectomy, Reconstruction (time to treatment completion only), Primary surgery, Surgery type, Postmastectomy immediate reconstruction type, Reconstruction /postmastectomy immediate reconstruction, Surgery decision, surveillance mammogram, in situ on index and pre index |
| Surgical outcomes/Side-effects/symptoms | Margins, Pathologic complete response, wound complication |
| Treatment duration | Prolonged treatment duration and treatment duration |
| Therapy type | Chemotherapy, Immunotherapy, Hormone therapy, Neoadjuvant Therapy, Radiotherapy |
| Trust in physician | Trust in surgeon |
| Tumor characteristics | Hormone receptors, ER/PR, HER2, Triple negative, Pathologic complete response, Tumor size, Tumor histology, TNM, Multifocal or multicentric tumor, Number of positive lymph nodes, or lymph node status/lymphovascular, lymphovascular invasion, N-stage, Clinical presentation, Grade, Breast cancer subtype/molecular subtype |
| Year of diagnosis |  |

### Study characteristics

A list of included studies and their key characteristics is provided in Table 2. Of the 36 studies, 12 indicated that examining disparity was a primary objective. Studies categorized ethnoracial groups in a variety of ways, mainly to support comparing Black and White populations [19,25–31]. Several studies also incorporated Hispanic and non-Hispanic ethnicity [32,33]. Many studies examined joint racial/ethnic categories, considering combinations of racial groups with Hispanic and non-Hispanic characteristics separately, such as Black non-Hispanic, in their analysis of race/ethnicity [33–42]. All included studies met the criteria for low risk of bias according to the CLARITY tool.

**Table 2.**
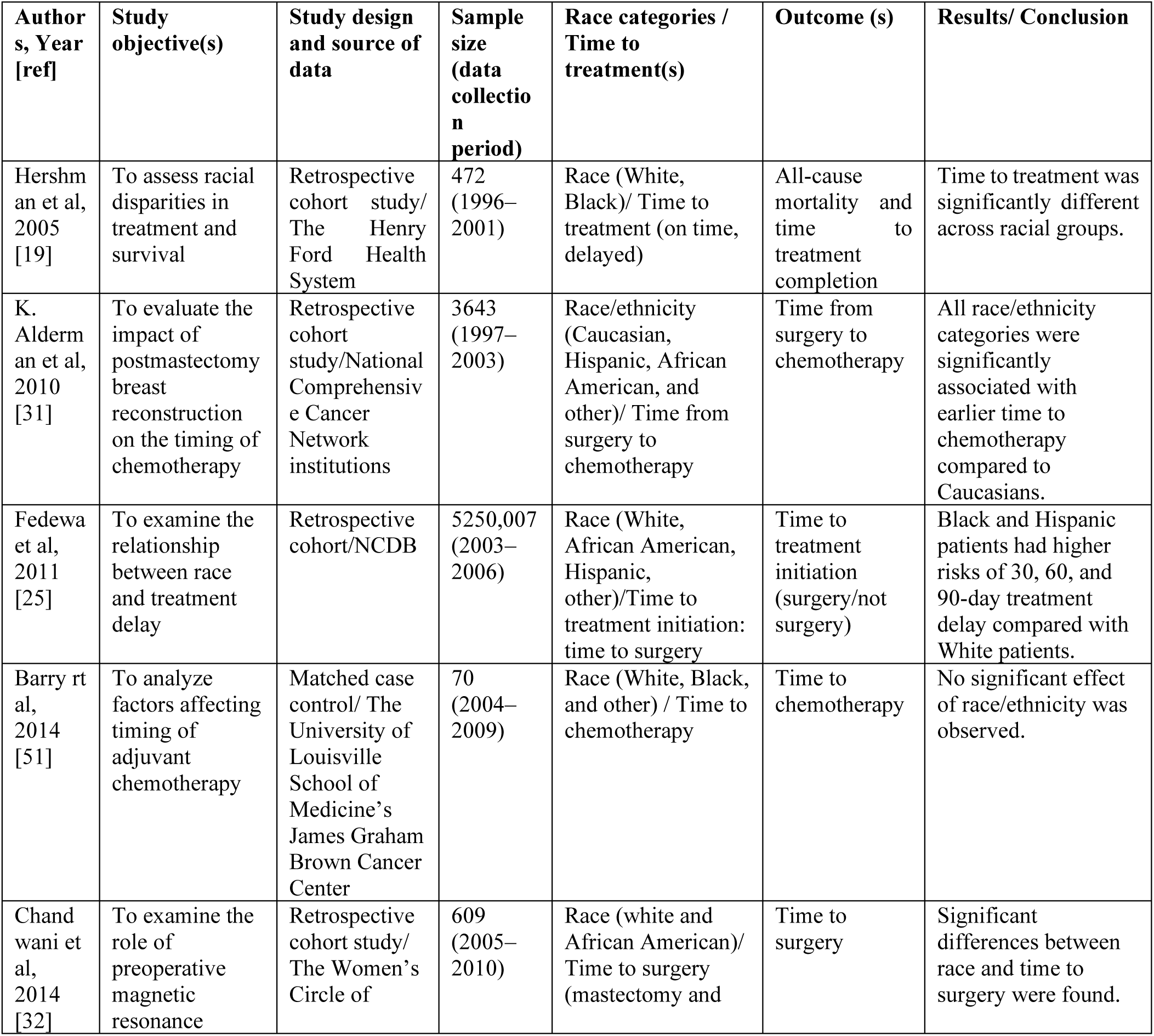
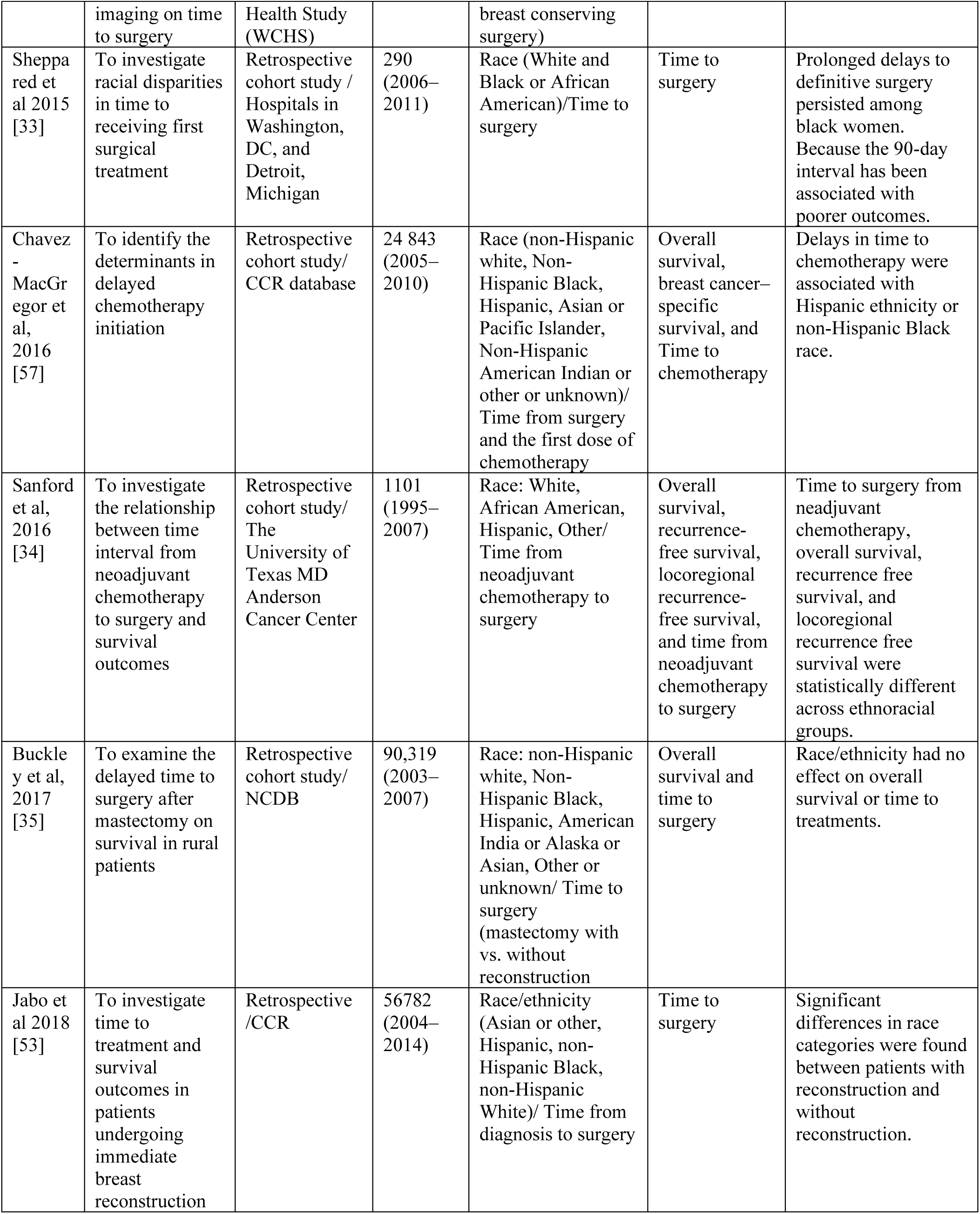
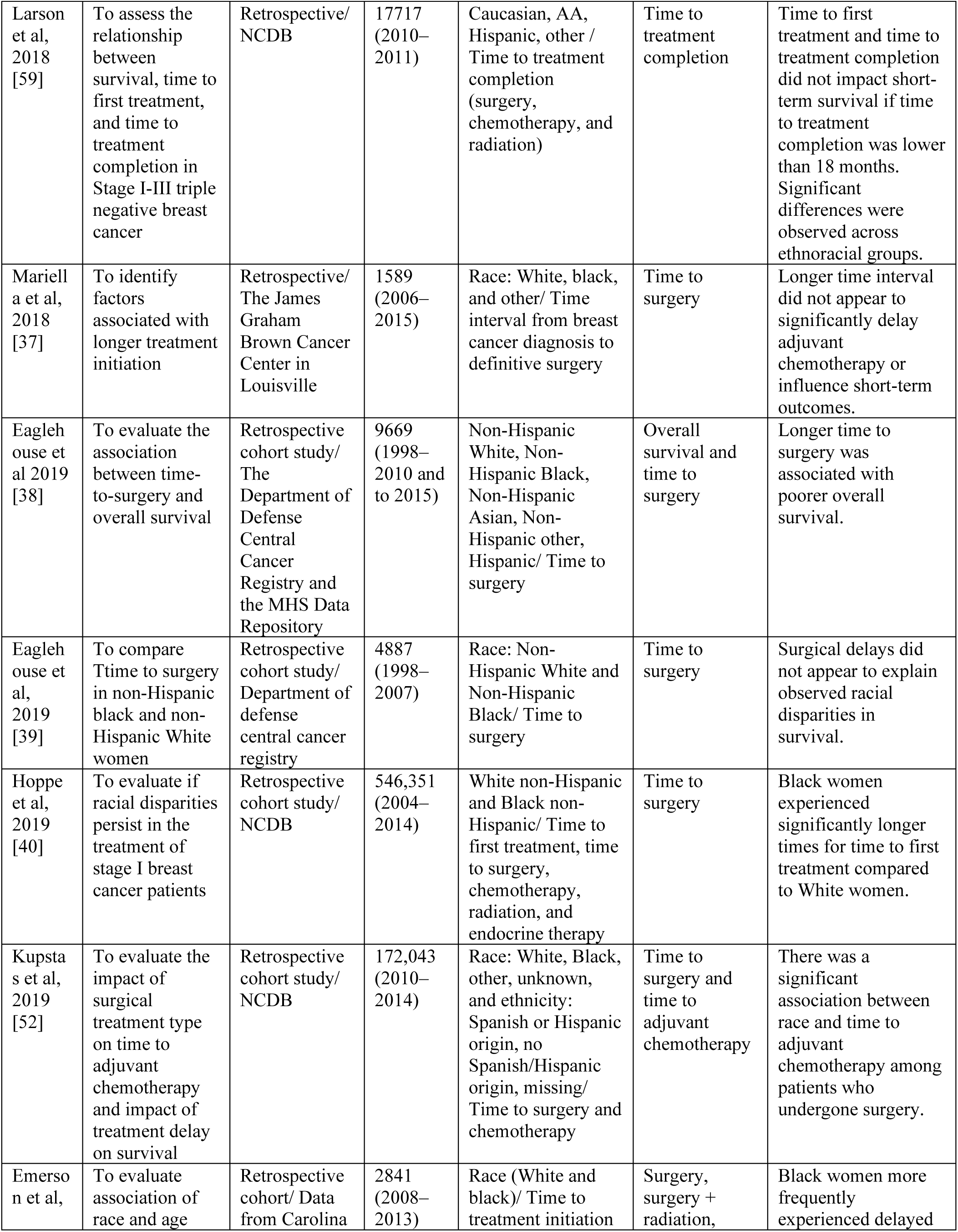
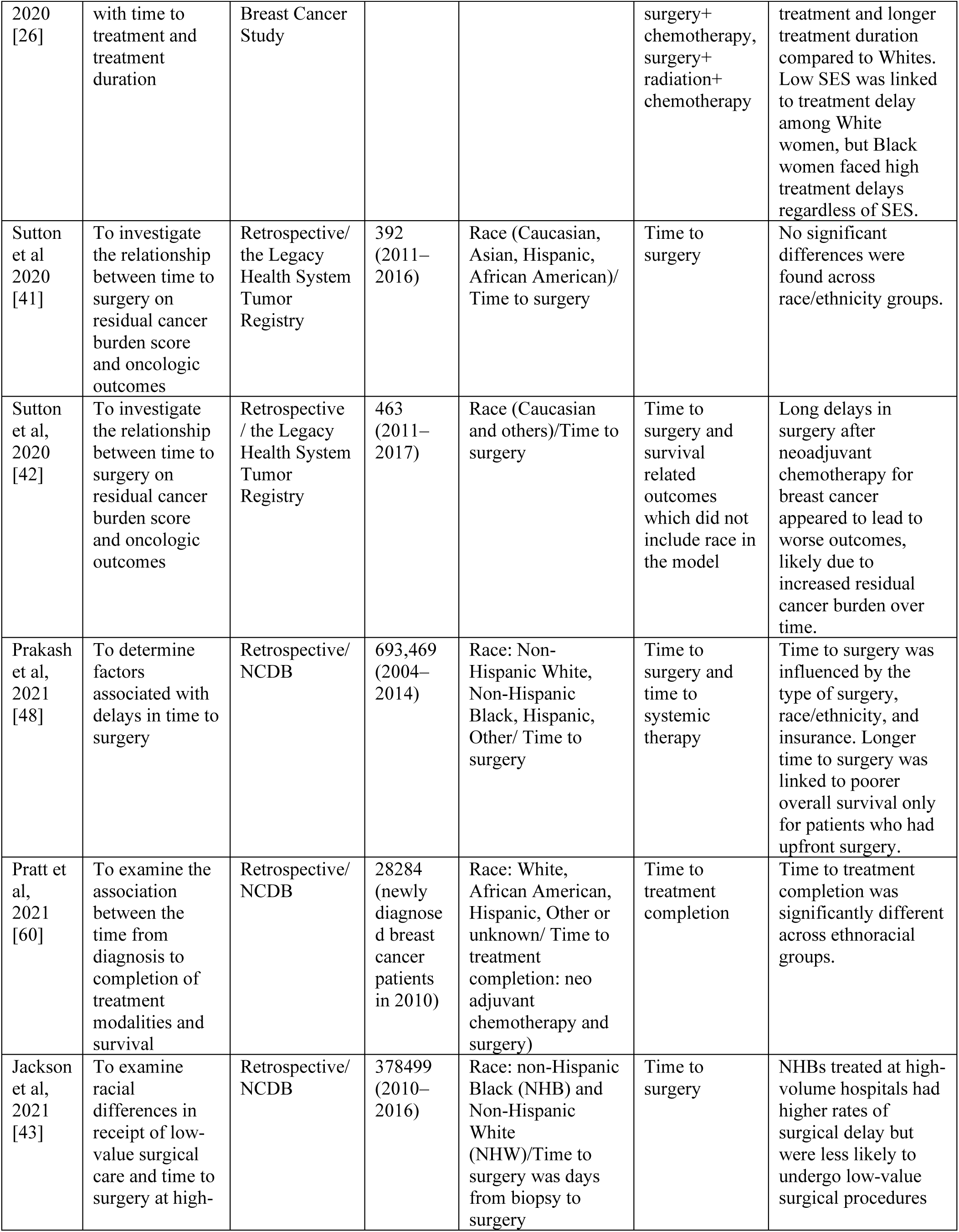
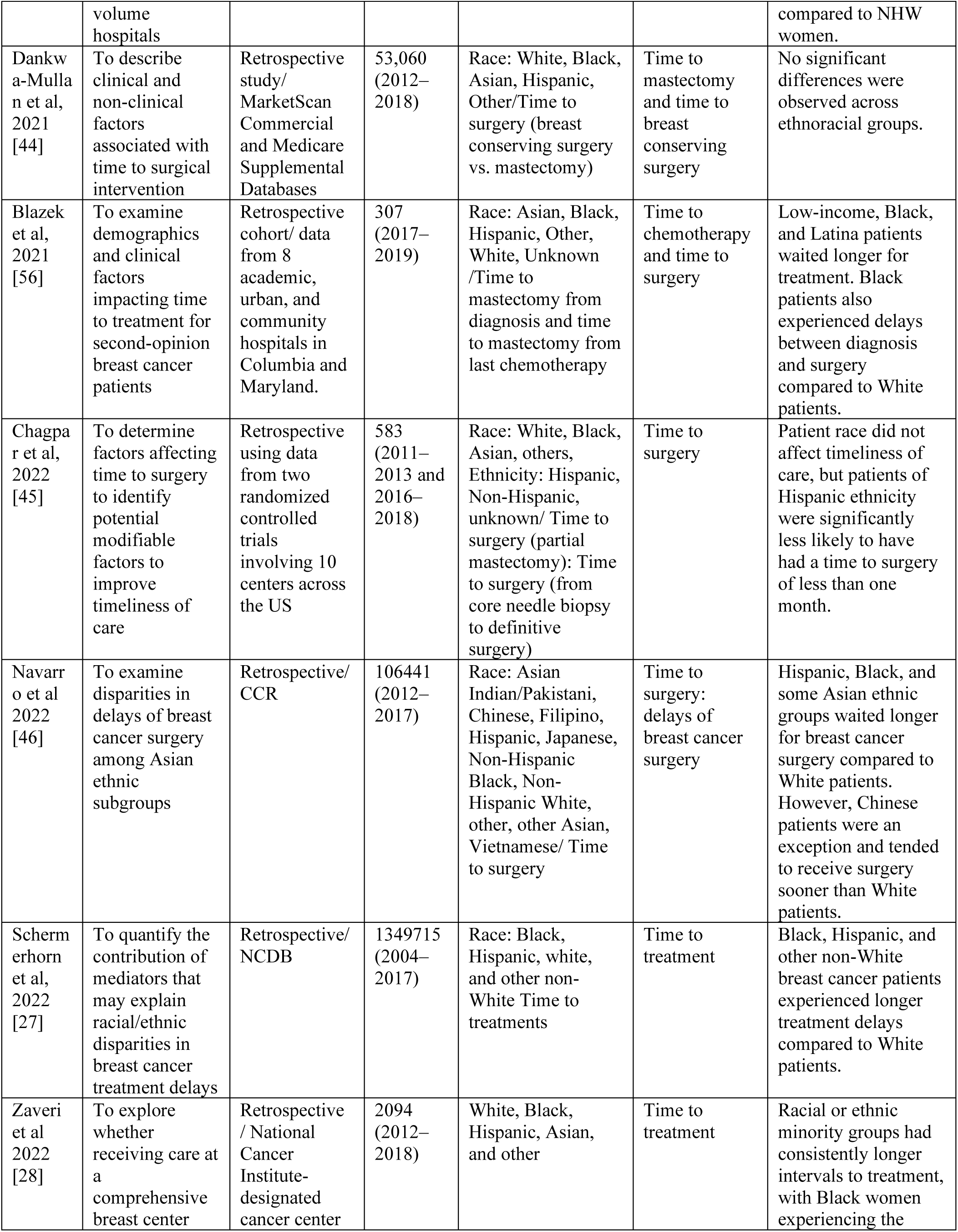
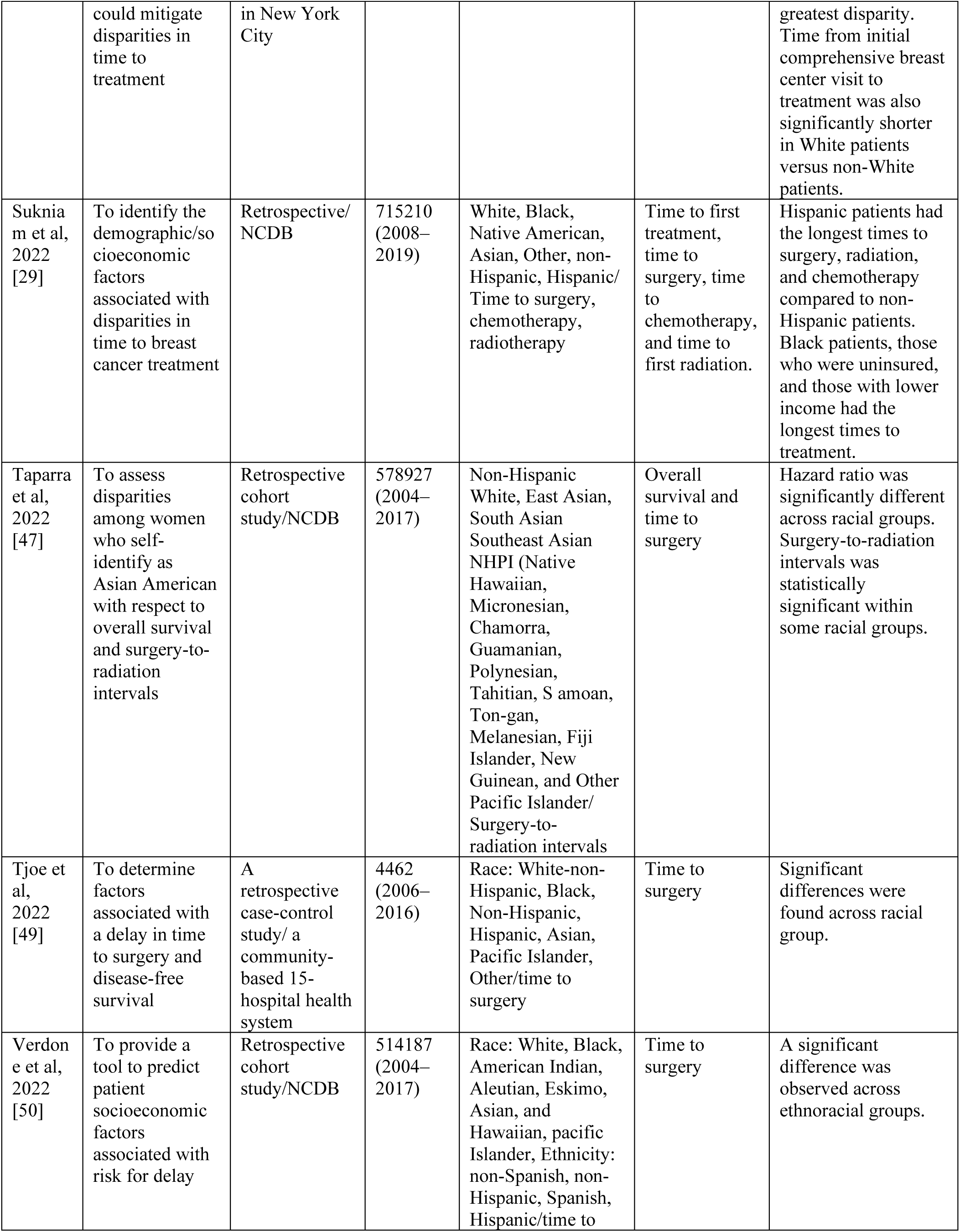
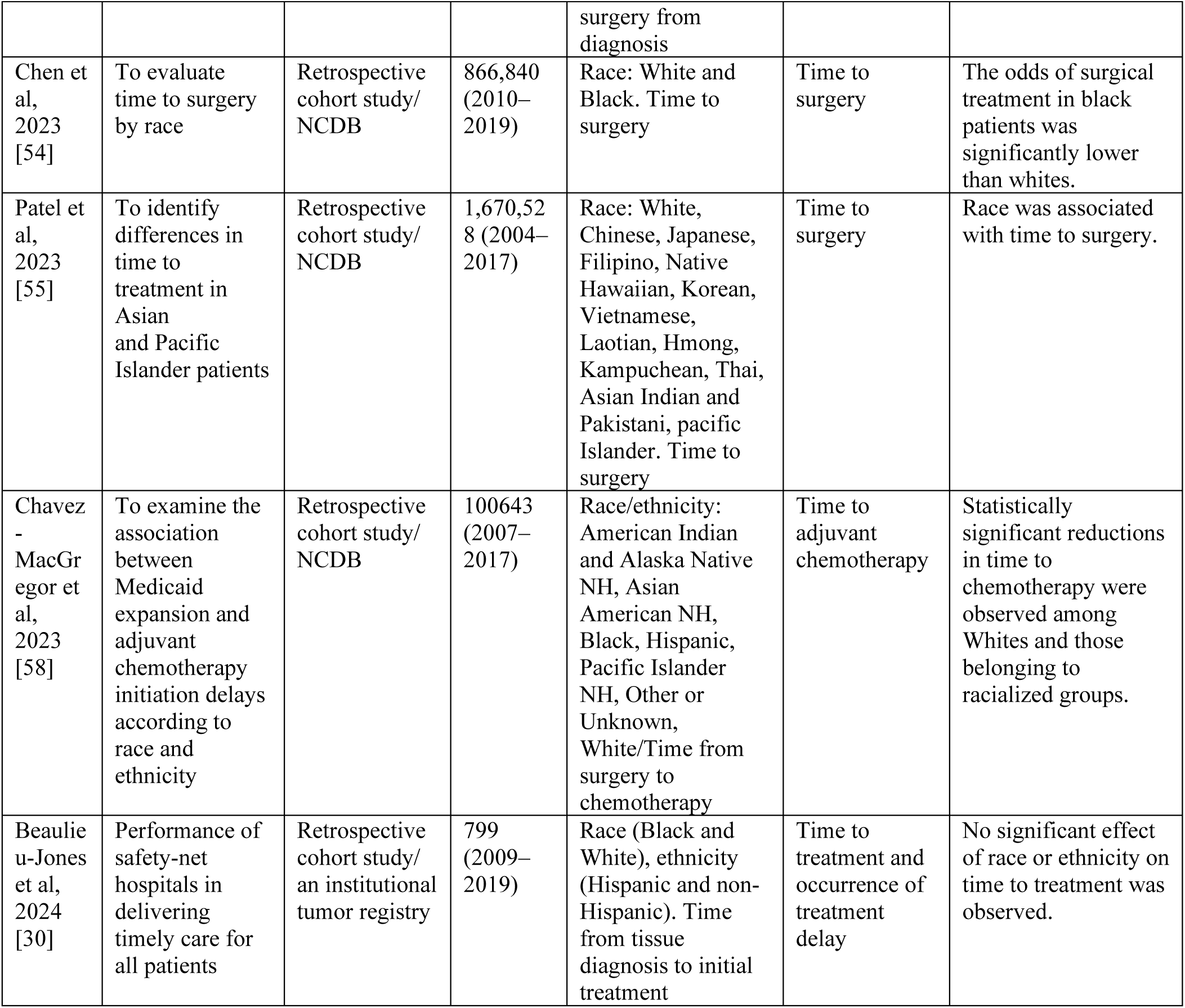
Descriptive characteristics of publications on time to treatment initiation across ethnoracial groups (NCDB: National Cancer Database, CCR: California Cancer Registry).

### Time to treatment initiation

The included studies used various measures of treatment initiation [19, 25–60]. These included time to treatment initiation (i.e., first treatment), time to treatment completion, and specific treatment of interest such as surgery and chemotherapy. Figure 2 shows the comprehensive causal DAG of the factors examined by these studies on the implied causal pathways to time to treatment and survival across different ethnoracial groups.

**Figure 2.**
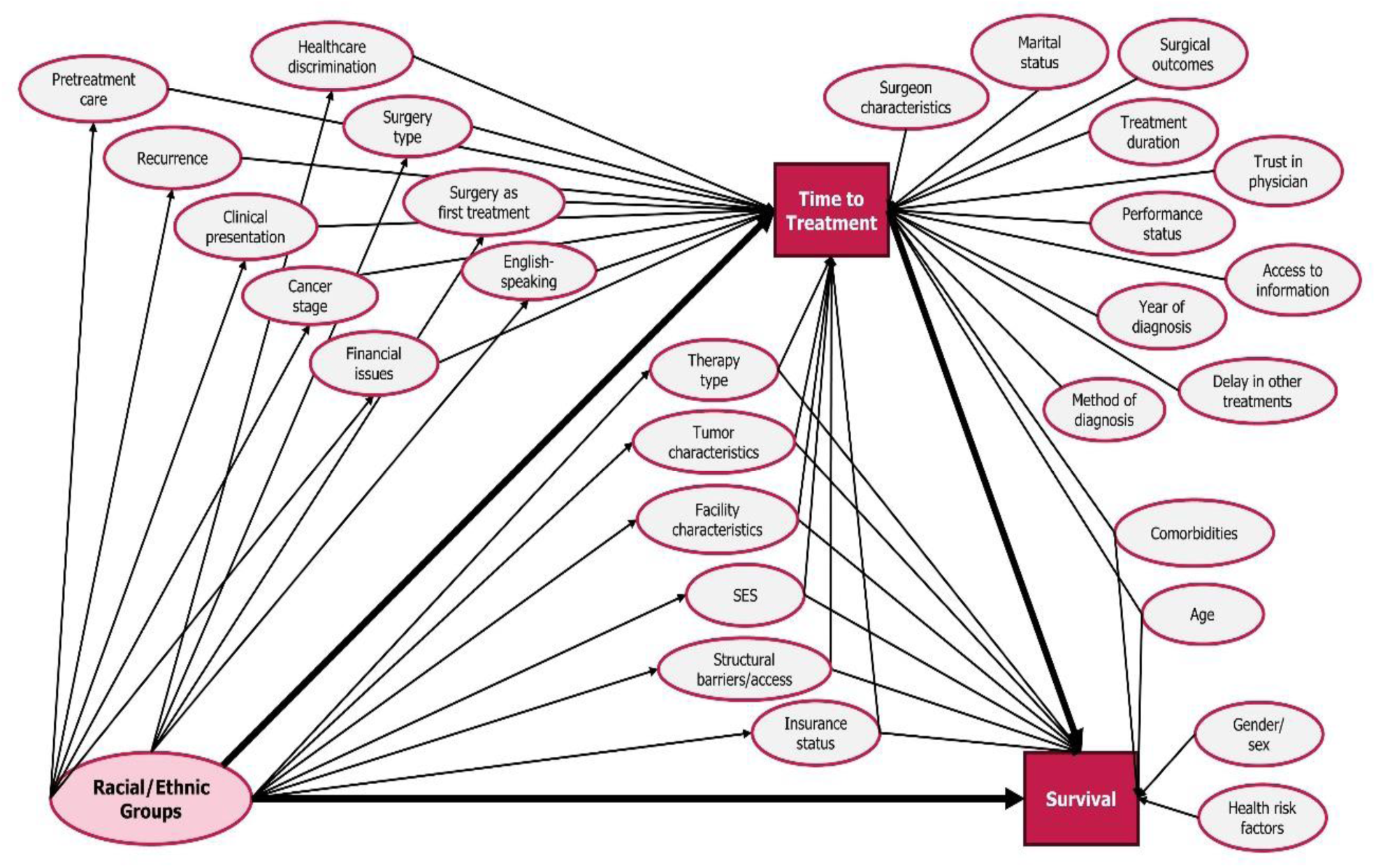
A composite causal DAG: Factors affecting time to breast cancer treatments and survival across racial/ethnic groups, as conceptualized in current literature.

#### i. Delay in time to first treatment

Five studies focused on time to initiation of any type of treatment in breast cancer patients [25–30]. Studies on time to first treatment found higher risk of delay in Black, Hispanic or other non-white races/ethnicities compared to White patients [25–27]. One study on delays in treatment initiation and treatment duration found that delayed initiation among Black patients was not significantly associated with either SES or access to care; however, low SES and more barriers were linked to longer treatment duration across Black and White groups [26].

#### ii. Delay in time to surgery

Time to surgery was specifically assessed in many studies, with most considering a wait of over one month as a delay in initiation of assigned surgery [31–55]. Mean time to definitive breast cancer surgery was significantly longer for Black patients than White patients [33,40]. Comparing factors across non-Hispanic Black and non-Hispanic White groups showed that Black patients treated at high-volume hospitals had higher rates of surgical delay but were less likely to undergo low-value surgical procedures compared to White patients [43]. A comparative analysis of this study showed significant disparities in the time to surgical treatment for breast cancer among Hispanic, non-Hispanic Black, and minority Asian ethnic subgroups compared to non-Hispanic White patients [43]. One study suggested that treatment delays among Black, Hispanic, and other non-white patients were explained mostly by disparities in education, comorbidities, insurance, and facility type [40]. Another study found the timeliness of care was not influenced by patient race; however, Hispanic patients were significantly less likely to undergo surgery within one month [44,51]. While immediate breast reconstruction delays time to definitive surgery, its use did not substantially affect time to adjuvant treatment or survival outcomes [51].

#### iii. Delay in time to chemotherapy

Studies addressed factors affecting the timing of chemotherapy in patients who are candidates for surgery [48,51,52,54,58]. Although no significant difference was found across ethnoracial groups, mastectomy with immediate reconstruction in breast-conservation candidates independently predicted delay in initiation of adjuvant chemotherapy [51]. One study indicated a significant difference in the delay of chemotherapy initiation across ethnoracial groups, with adverse outcomes linked to a delay in initiating adjuvant chemotherapy of three months or more [53].

### Ethnoracial disparities, time to treatment and survival

Seven addressed both time to treatment and survival together [19, 34,36,39,47,52,53, 56–59]. A large retrospective study that assessed disparities in Asian Americans with Native Hawaiians and Other Pacific Islanders (NHPI) found NHPI had worse survival compared to non-Hispanic Whites, while all Asian American subpopulations had improved survival, with Southeast Asians and NHPI experiencing longer times to treatment initiation [47]. One study found that longer time to surgery correlates with poorer overall survival in breast cancer across different ethnoracial categories, after adjusting for time to treatment [39].

### Comprehensive Summary of Causal Pathways

The comprehensive DAG in Figure 2 includes all assessed factors affecting time to treatment and survival in breast cancer patients and their relationships, providing a visual summary of the causal conceptualization in the summarized literature. Note that while the comprehensive DAG shows many mediated pathways, these were often constructed of implied pathways from multiple studies that included portions of the pathway.

### Study author-identified unmeasured factors affecting TTI and Survival

Most included studies used administrative records of cancer registry programs, with only one undertaking primary data collection. Given limitations of secondary data sets, authors highlighted numerous unmeasured factors that could significantly impact the initiation of treatment in breast cancer patients; these are summarized chronologically in Table 3. These change over time; for instance, an earlier study identified additional imaging or biopsy procedures as key missing factors affecting the time to treatment, and subsequent research showed that these factors exhibit variations across different socioeconomic status (SES) levels, ultimately associated with ethnoracial groups [11,12].

**Table 3.**
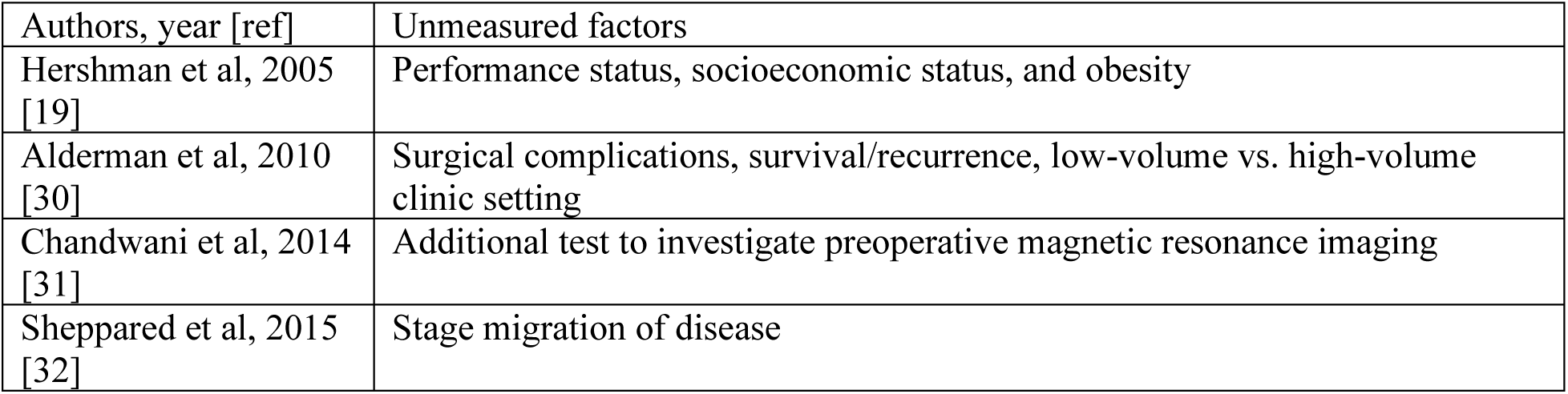
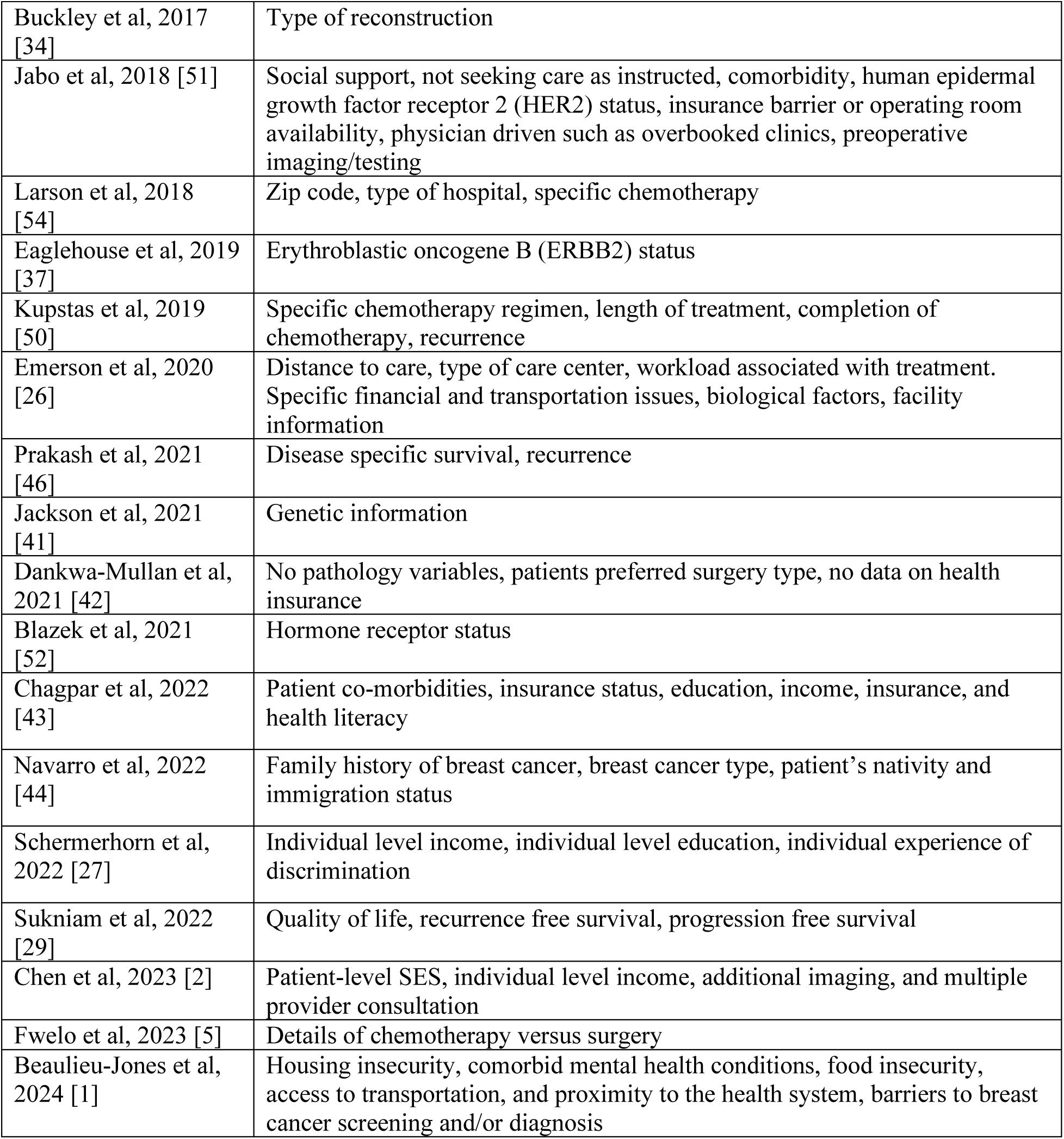
List of unmeasured factors specified by authors in include studies.

In addition to factors that authors of included studies identified as unmeasured or needed, we drew on knowledge of ethnoracial disparities research to identify additional explanatory factors that were missing from the comprehensive DAG, and thus from the literature, particularly those at higher (e.g., structural) social levels rather than the individual level. These included factors related to health systems, and historic and structural racism. To illustrate these, we drew on social ecological models to create a new biosocioecological model of ethnoracial disparities, time to treatment, and survival (Figure 3) [61]. Factors were grouped within four levels: tumor, individual, health system, and society, and were identified from both included studies, and from additional theory and literature on US ethnoracial disparities.

**Figure 3.**
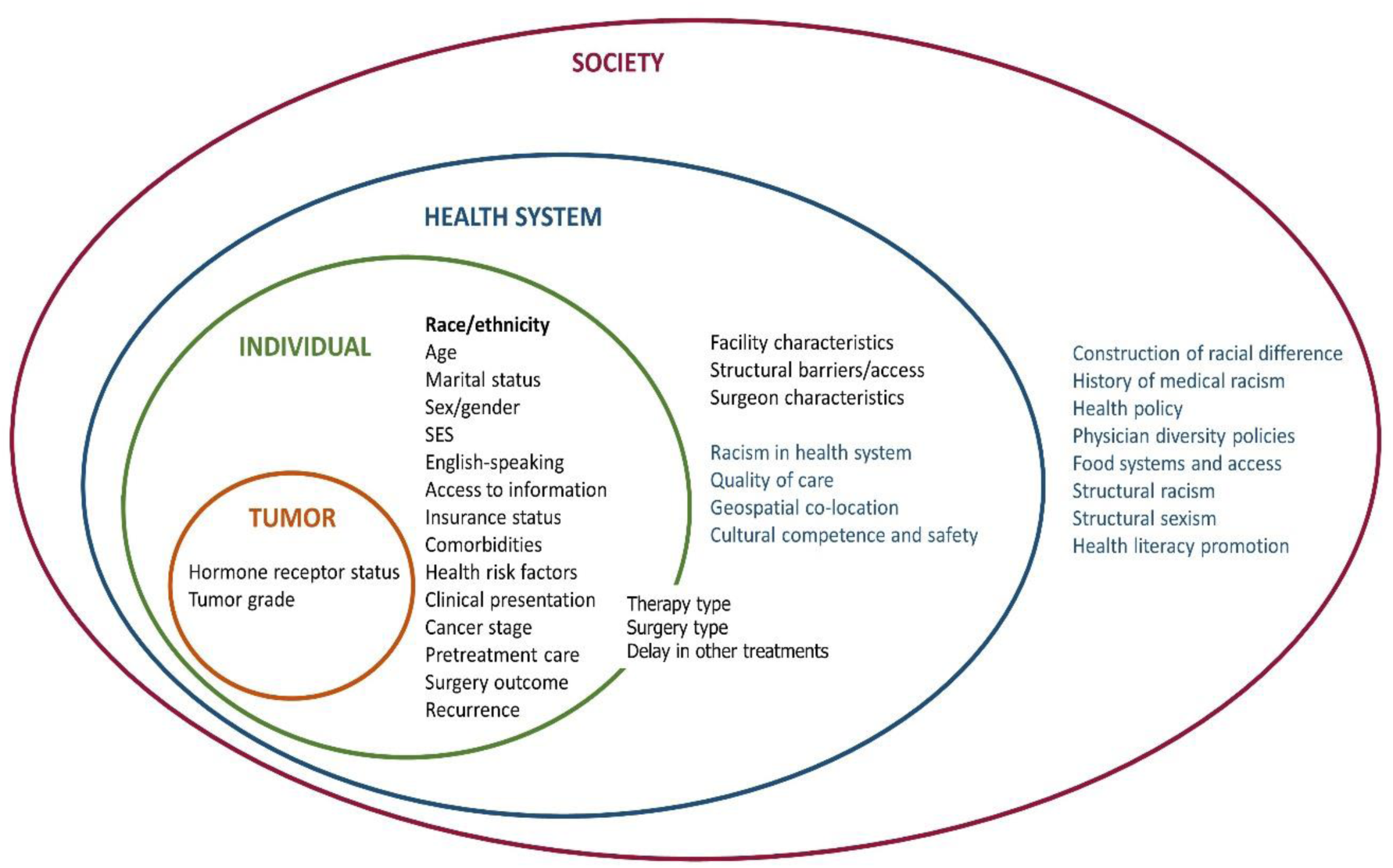
A biosocioecological model on racial and/or ethnic disparities and time to treatment and survival in breast cancer, including factors identified in a systematic review (black) and disparity-related factors not found in review (blue)

## Discussion

Ethnoracial disparities in the timing of breast cancer treatment initiation and completion are clear, with delays significantly impacting survival in patients [27,32,39,42,46,47]. Other factors are also clearly associated with delay; for example, older patients tend to have more delay than younger individuals [22,43,47,54], possibly in response to therapy type, healthcare access, and patient preferences. At the individual patient level, these disparities highlight the importance of improving surgeon-patient communication, decision-making, and care coordination throughout neoadjuvant systemic therapy and the perioperative period [36]. At the system level, they highlight the need for a nuanced understanding of the driving factors of delays and their causal relationships (as expressed in the DAGs) and structural barriers (as expressed in the biosocioecological model) to ensure that system-level interventions to reduce delays are effective.

### Causal interpretation of race/ethnicity in the literature

The causal interpretation of race/ethnicity is challenging [62]. Studies mainly investigated the association of time to treatment initiation and survival, controlling for confounders such as age and stage, across ethnoracial groups. This may be because many factors that impact ethnoracial inequities in health are rarely measured in breast cancer studies and do not typically exist in registry data, for example, racist experiences in health systems and SES early in life. The persistence of not measuring important causal factors and not attending to causal considerations within the context of ethnoracial health disparities in breast cancer remains a concern, because their absence impedes our ability to develop and assess interventions designed to redress such disparities.

### Focus on individual versus structural factors

Although studies frequently identified unmeasured factors that were deemed important, (described in Table 3), there was an exclusive focus on individual-level rather than structural factors that lead to delays in treatment and poorer survival. For example, studies did not measure or examine the relationship between structural racism, structural sexism, and their intersections. Neither did studies examine the interaction between health system-level factors, such as quality of care, and individual-level factors such as SES. This lack of consideration for broader structural influences limits our understanding of the complex interplay between tumor, societal, healthcare system, and individual-level factors in shaping breast cancer outcomes. One large cohort study specifically discussing system-level factors (i.e., hospital level), found that there were significant differences in time to surgery for Black women than White women among nonmetastatic invasive breast cancer patients. This difference varied across different hospitals and regions in the country, suggesting that hospital factors play a significant role in disparities in surgical care. However, studied system-level properties were considered as individual level factors in the analysis [54].

### A biosocioecological model for breast cancer disparity

The lack of a system-level lens in this area led us to consider how disparity has been conceptualized in this field and to consider what drivers of disparity, both individual and structural, may be missing from this conceptualization. In response, we have created a biosocioecological model to describe factors across different social levels that may impact breast cancer disease progression and survival. The existence of a substantial number of factors in this model, despite not being included in the earlier survey research, demonstrates a significant gap in the body of knowledge regarding disparities in breast cancer treatment and survival. This observation underscores the predominant focus of research on breast cancer at the individual level. While individual factors undoubtedly play a significant role in shaping breast cancer outcomes, the emphasis on higher-level systemic and societal influences is essential for comprehensively understanding the complexities of this disease. By recognizing the broader structural determinants that impact breast cancer incidence, treatment access, and outcomes, researchers can develop more holistic approaches that address the full spectrum of factors contributing to ethnoracial disparities in breast cancer.

### Limitations

We only set out to retrieve documents in English, and given our exclusively US-based results, our search strategy may have inadvertently excluded work in other jurisdictions where the discourse on ethnoracial disparity is framed differently. We also excluded gray literature, which may have excluded community and popular perceptions of such disparities in breast cancer. Future work that explicitly seeks a more global and non-academic perspective may further our understanding. Additionally, the absence of causal designs in studies, for example, using matching, propensity scores, identification, and control of confounders, as well as the lack of mediation models, limit our ability to assess with certainty what the causal assumptions of the authors might have been. Although some studies implied the presence of mediation models, there was a lack of formal mediation analysis in this context.

## Conclusion

There are persistent disparities observed in treatment and mortality outcomes among different ethnoracial groups, particularly between White and Black female breast cancer patients. This highlights the importance of addressing and eliminating ethnoracial disparities in breast cancer care to ensure equitable access to timely and effective treatments for all patients, ultimately improving long-term survival rates across diverse populations. Future research must adopt a more inclusive and multi-level approach, incorporating systemic and societal factors alongside individual-level determinants to advance our understanding and improve interventions in breast cancer care.

## Data Availability

Not applicable

## Authors’ contributions

PMH initially conceptualized the project. PMH, DJL and GRB critically reviewed the methodology and the content, and designed the study protocol. PMH and DJL performed data extraction and risk of bias assessment. All authors assisted with the construction of the composite DAG and conceptualizing the biosocioecological model. PMH wrote the first draft of the manuscript. The final manuscript was edited and approved by all authors.

## Acknowledgements

We would like to thank Lux Li for his help with screening full texts and initial data extraction, and Wanlin Hu for review of an earlier draft.

